# Senescence-related cytokine levels are associated with HIV-1 serostatus and persistence

**DOI:** 10.1101/2025.02.05.25321757

**Authors:** Yijia Li, Zoamy N. Soto-Ramirez, Jenny Roscher, Tom Medvec, Mounia Alaoui-El-Azher, Paolo Piazza, Yue Chen, Nicolas Sluis-Cremer, Charles Rinaldo, Bernard JC Macatangay

## Abstract

**Background:** HIV-1 is associated with accelerated aging. The senescence-associated secretory phenotype (SASP) includes biological and cytokine profiles that induce cellular senescence and inflammaging. In this study, we leveraged the Multicenter AIDS Cohort Study (MACS) to evaluate the role of SASP in aging, HIV-1 reservoir, and inflammation in people with HIV-1 (PWH) on long-term suppressive antiretroviral therapy (ART).

**Methods:** In this retrospective study we included plasma and serum samples from 27 virally-suppressed PWH and 10 people without HIV-1 (PWoH) collected in 2019 and 2023. SASP markers were quantified in the 2019 and 2023 samples. Plasma residual viremia, intact and defective proviral DNA were quantified in the 2019 samples. Correlations between SASP markers and HIV-1 reservoir were performed using the Spearman test, and the sparse partial least squares discrimination analysis was used to identify variables that distinguish HIV-1 serostatus.

**Results:** All study participants were male with a median age of 59 years. SASP markers did not show significant changes longitudinally in either group. We identified a set of markers that had moderate performance in distinguishing PWH and PWoH, including CMV serum antibody titer, matrix metalloproteinase 9 (MMP-9), Growth/differentiation factor-15, Stanniocalcin-1 and SerpinE1. Among all the SASP markers, MMP-9 was significantly associated with intact HIV-1 proviral levels (rho=0.60, P=0.002).

**Conclusion:** In this cohort study, we revealed the relationship between SASP markers and HIV-1 persistence. Future interventions targeting the senescence pathways may impact HIV-1 persistence.

## Introduction

Non-AIDS complications, including cardiovascular diseases, chronic renal and liver diseases etc., significantly contribute to morbidity and mortality in people with HIV-1 (PWH) including those on suppressive antiretroviral therapy (ART) [1]. ART is not curative and HIV-1 persists indefinitely in most PWH [2]. Immunosenescence and aging-related inflammation (inflammaging) has emerged as a significant underlying mechanism associated with chronic diseases [3], and HIV-1 is thought to significantly contribute to immunosenescence [4] and accelerated aging [5].

Senescence-associated secretory phenotype (SASP), a group of inflammation and immune regulators, are key mediators in aging [6]. SASP is secreted by senescent cells to trigger proinflammatory responses and tissue repair via immune-mediated elimination of the senescent cells; however, they are not always effective and may drive chronic inflammation and immune dysfunction [6]. The relationship between immunosenescence and measures of HIV-1 persistence is poorly understood. To this end, we leveraged the Multicenter AIDS Cohort Study (MACS, now part of the MACS/ Women’s Interagency HIV-1 Study Combined Cohort Study, MWCCS) to interrogate this knowledge gap.

## Methods

### Participants and sample collection

Peripheral blood mononuclear cells (PBMC) and plasma samples were collected from 27 male PWH on ART in the Pittsburgh Clinical Research Site of the MWCCS from 2019 and 2023 visits. Participants were enrolled if they had an HIV-1 viral load <30 copies/ml for at least two years and a CD4+ T cell count >400 cells/mm^3^. Ten male Pittsburgh MWCCS participants without HIV- 1 with matched age were included as a control group. All participants provided written informed consents. This study is consistent with the Declaration of Helsinki.

### Integrase single copy assay (iSCA)

We used a quantitative RT-PCR assay with single-copy sensitivity targeting HIV-1- 1 *Integrase* (*IN*) RNA to quantify plasma viremia, as described previously [7].

### Quantification of SASP soluble markers

We quantified endothelial plasminogen activator inhibitor (Serpin-E1), matrix metalloproteinase 2 (MMP-2), matrix metalloproteinase 9 (MMP-9), interleukin-1 receptor antagonist (IL-1RA), growth/differentiation factor 15 (GDF-15), and Stanniocalcin-1 (STC-1) using the FlexMAP 3D Luminex platform (Thermo Fisher Scientific, Waltham, MA, USA) with Luminex Discovery Assay - Human Premixed Multi-Analyte Kit (R&D Systems, Minneapolis, MN, USA) per manufacturers’ instructions.

### Intact proviral DNA assay (IPDA™) measurement

CD4+ T cells were purified from 1 x 10^7^ cryopreserved PBMC using the EasySep Human CD4+ Isolation kit (Stem Cell Technologies, Seattle, WA, USA). After magnetic isolation, 95% purity of the CD4+ T cells was verified by flow cytometry. Genomic DNA was then extracted with the QIAamp DNA Mini Kit (Qiagen, Germantown, MD, USA). DNA concentrations were determined using the Qubit 3.0 and Qubit dsDNA BR Assay Kit (Thermo Fisher Scientific, Waltham, MA, USA). Integrated proviral DNA was measured using a published procedure [8]. After correction for DNA shearing, results were expressed as integrated proviral copies per 1 x 10^6^ CD4+ T cells.

### Cytomegalovirus (CMV) serum antibody (Ab) measurement

Serum CMV immunoglobulin G (IgG) Ab levels were measured using an ELISA-based assay (EUROIMMUN, Lübeck, Germany) per manufacturer’s instructions.

### Statistical analysis

Continuous variables were analyzed using non-parametric tests including Wilcoxon signed-rank test (if independent) and Wilcoxon rank-sum test (if paired). Categorical variables were analyzed using Chi-squared test or Fisher exact test. P<0.05 was deemed statistically significant unless stated otherwise. Geometric means of all SASP markers are reported in the log_10_ form and calculated using

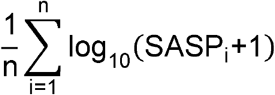

Sparse Partial Least Squares Discriminant Analysis (sPLS-DA) was performed with the “mixOmics” package (version 6.26.0) to distinguish PWH and PWoH using SASP markers and CMV Ab titers. We first determined the optimal number of components, and then the sPLS-DA model was tuned with 5-fold cross-validation 50 times, with n = 10 as the limitation of the maximum number of variables in each component. The optimal number of variables and number of components associated with the lowest balanced error rate were applied to the final models. Variable importance in projection (VIP) scores were determined to demonstrate the contribution of selected variables to the first component [9]. Statistical analyses were conducted in R (4.3.1) and Stata SE (Version 18).

## Results

Thirty-seven male participants were included in this analysis, including 27 PWH and 10 people without HIV-1 (PWoH). Median age (at the 2019 visit) was 59 years in both groups. Median CD4+ T cell count was 634 cells/mm^3^ at the time of 2019 visit (interquartile range [IQR], 548- 760 cells/mm^3^) and 690 cells/mm^3^ (IQR 542-951, available CD4+ T cell count in 2023 n=20, paired P=0.9); nadir CD4+ T cell count was 272 cells/mm^3^ (IQR 148-456 cells/mm^3^). All PWH had undetectable HIV-1 viral load by clinical laboratory testing at the 2019 and 2023 visits.

Other demographic characteristics and ART regimens are shown in Supplemental Table 1.

Neither the SASP markers nor CMV Ab titer differed significantly between PWH and PWoH, at either time point. The SASP markers did not change significantly over time, whereas the CMV Ab titers increased in both PWH (P=0.0002) and PWoH (P=0.014) (Supplementary Figure S1A and 1B). Using sPLS-DA, we could select a set of markers that distinguish PWH and PWoH. Using the 2019 dataset, CMV Ab, MMP-9 and GDF-15 were the top 3 most important variables to distinguish PWH and PWoH (Figure 1A and 1B). Using the 2023 dataset, CMV Ab, STC-1 and SerpinE1 were the top 3 most important variables (Figure 1C and 1D). Sensitivity analyses excluding CMV-seronegative participants yielded similar results (Supplementary Figure S2A- S2D).

**Figure 1.**
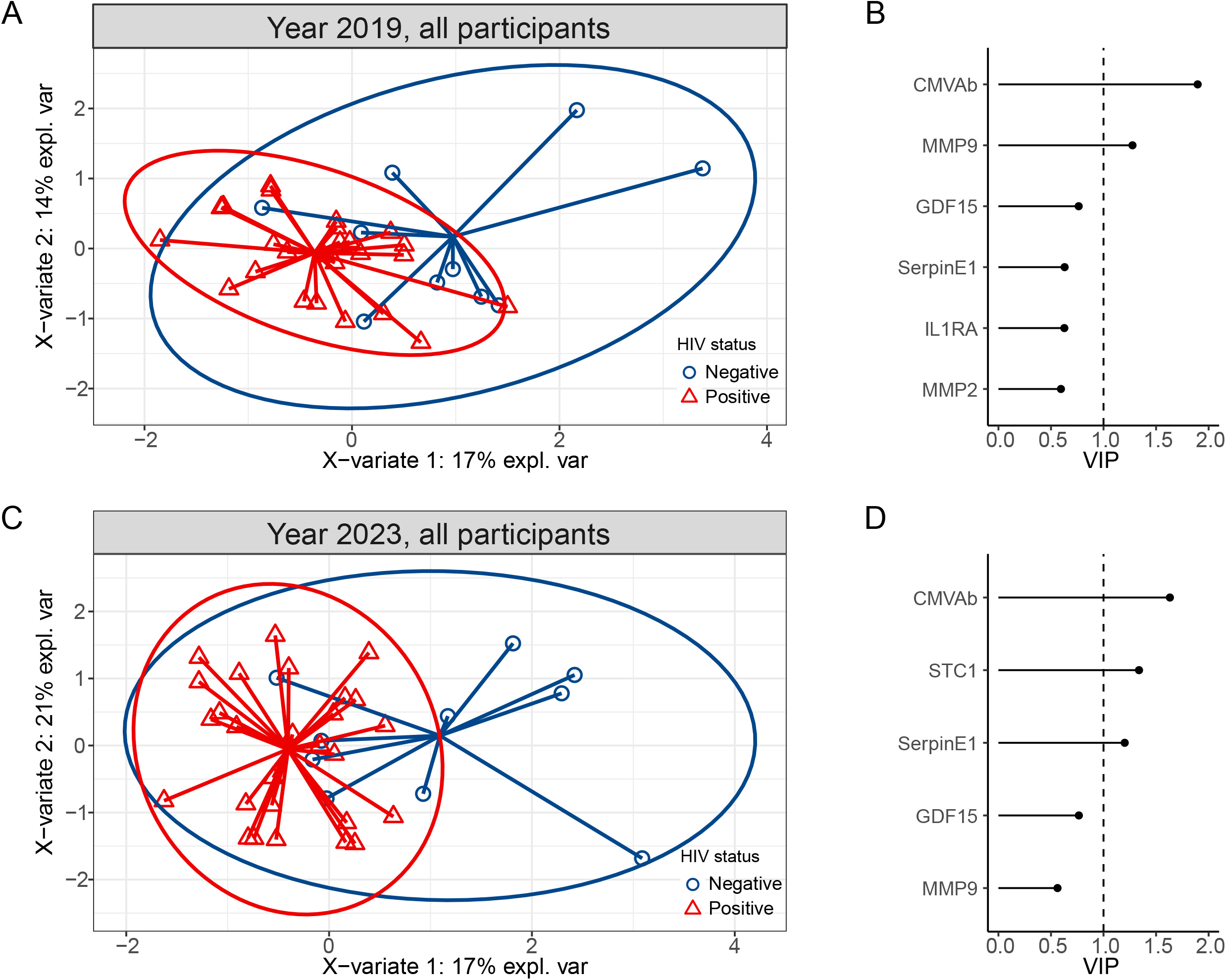
Partial least squares discriminant analysis (PLS-DA) to evaluate the association between SASP markers, CMV Ab, and HIV-1 serostatus. (A) SASP markers and CMV Ab from 2019; (B) Variable importance in projection (VIP) of selected SASP markers from 2019 that can distinguish HIV-1 serostatus. (C) SASP markers and CMV Ab from 2019; (D) Variable importance in projection (VIP) of selected SASP markers from 2019 that can distinguish HIV-1 serostatus.

Chronological age was significantly associated with levels of GDF-15 and IL1RA in 2019 (Figure 2A). MMP-9 was significantly associated with HIV-1 intact proviral levels (rho=0.60, P= 0.002; Figure 2A and 2B). Other SASP markers were not associated with HIV-1 intact, total, or defective proviral levels (Figure 2A). The geometric mean (GM) of all SASP markers, however, showed a moderate correlation with total (rho=0.47, P=0.02) and 5’ defective (rho=0.47, P=0.02) proviral levels, in addition to CMVAb titers (Figure 2A and 2C-2D).

**Figure 2.**
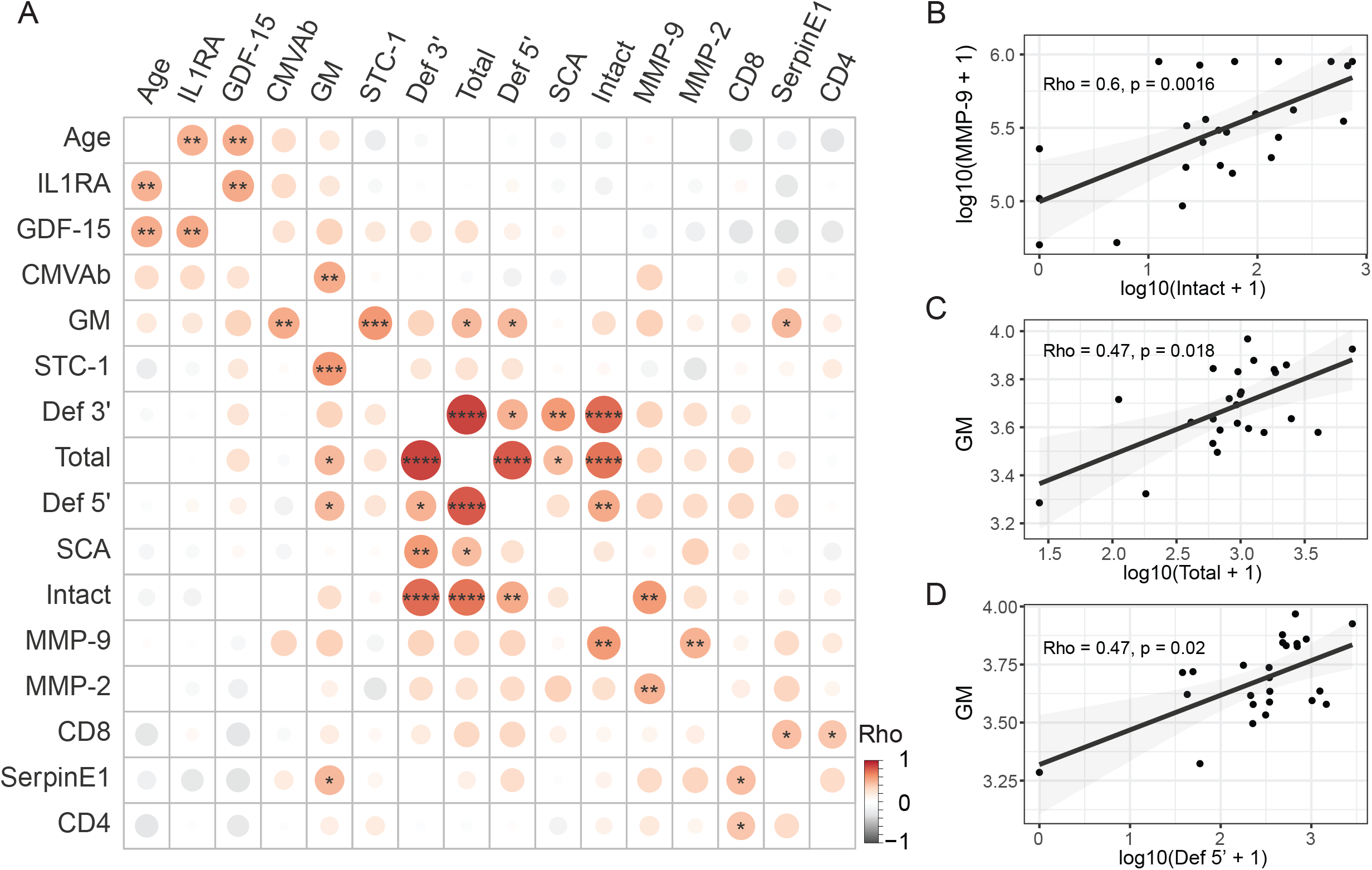
Correlations among SASP markers, CMV Ab, chronological age, and HIV-1-related parameters in all participants. (A) Heatmap showing Spearman correlations among variables.****, P<0.0001; ***, P<0.001; **, P<0.01; *, P<0.05. (B) Spearman correlation between intact HIV-1-1 proviral levels and MMP-9. (C) Spearman correlation between total HIV-1-1 proviral levels and geometric mean (logarithmic 10) of SASP markers from 2019. (D) Spearman correlation between 5’ Defective HIV-1-1 proviral levels and geometric mean (logarithmic 10) of SASP markers from 2019.

## Discussion

This study evaluated the relationship between inflammaging and measures of HIV-1 persistence. Inflammaging in PWH is reported to promote premature and accelerated aging at molecular and functional levels, as well as HIV-1-related non-AIDS comorbidities [5,10]. In this study, we demonstrate that the SASP markers MMP-9, GDF-15, STC-1 and SerpinE1, along with CMV Ab, can distinguish PWH from PWoH at comparable age, which further emphasizes the association between HIV-1 and inflammaging.

In our assessment of inflammaging and HIV-1 persistence, we identified a significant association between MMP-9 and intact HIV-1 proviral levels. To our knowledge, this association has not been previously described. Of note, MMP-9 plays a crucial role in the extracellular matrix remodeling process, which involves the breakdown and reorganization of proteins and other components that support and connect cells. A role for MMP-9 has also been implicated in several HIV-1-associated comorbidities including HIV-1-associated neurological disorders (HAND) [11], liver diseases [12], and lung diseases [13], as well as a damaged gastrointestinal barrier [14] associated with systemic immune activation and other HIV-1-related comorbidities [15]. MMP-9 may also promote clonal expansion of long-lived CD4+ T cells that harbor intact HIV-1 provirus: although it has not been reported in HIV-1 infection, MMP-9-expressing macrophages in the breast cancer microenvironment positively correlate with T cell expansion after anti-programmed cell death protein-1 (PD-1) treatment [16]. In addition, MMP-9 secretion by HIV-1-infected monocytes promotes viral dissemination [17], although this is less likely to directly contribute to an increase in intact proviral levels. Further studies evaluating the underlying mechanisms are needed.

Our results support an association between CMV Ab titers and HIV seropositivity, which has been previously identified [18,19]. We also see an increase in CMVAb titer with time. These observations presumably represent periodic reactivation of latent, persistent CMV infection in PWH that creates an inflammatory milieu[20], likened to SASPs driving aging in PWH in our study.

We recognize limitations in our study. The sample size was relatively limited (PWH n=27 and PWoH n=10) and all participants were male assigned at birth. Furthermore, we only evaluated this association at one time point (the year 2019). Expanding the analyses to a larger sample size, including female participants and multiple time points, should provide further insight.

In conclusion, our study supports that HIV-1 infection is associated with changes in SASP markers and elevation in CMV Ab titers. HIV-1 intact proviral levels are specifically associated with MMP-9. Additional studies evaluating the mechanisms linking SASP, HIV-1 persistence, and CMV infection are warranted.

## Data Availability

All data produced in the present work are contained in the manuscript.

## Acknowledgment

The authors gratefully acknowledge the contributions of the study participants and the dedication of the staff at the University of Pittsburgh Clinical Research Site (CRS). Data in this manuscript were collected by the University of Pittsburgh CRS of the MACS/WIHS Combined Cohort Study (MWCCS).

## Notes

### Competing Interest Statement

This study was funded in part by the National Heart, Lung, and Blood Institute (U01-HL146208), the MWCCS Data Analysis and Coordination Center (U01‐HL146193), and the Rustbelt Center for AIDS Research (P30 AI036219), together with internal funding from the Department of Medicine, University of Pittsburgh School of Medicine. The contents of this publication are solely the responsibility of the authors and do not represent the official views of the National Institutes of Health (NIH).

### Author Declarations

This study is approved by University of Pittsburgh IRB.

## Reference

1. Trickey A, Ambia J, Glaubius R, et al. Excess mortality attributable to AIDS among people living with HIV in high-income countries: a systematic review and meta-analysis. J Int AIDS Soc 2024; 27:e26384.

2. Churchill MJ, Deeks SG, Margolis DM, Siliciano RF, Swanstrom R. HIV reservoirs: what, where and how to target them. Nat Rev Microbiol 2016; 14:55–60.

3. Liberale L, Badimon L, Montecucco F, Lüscher TF, Libby P, Camici GG. Inflammation, Aging and Cardiovascular Disease: JACC Review Topic of the Week. J Am Coll Cardiol 2022; 79:837–847.

4. Lagathu C, Cossarizza A, Béréziat V, Nasi M, Capeau J, Pinti M. Basic science and pathogenesis of ageing with HIV: potential mechanisms and biomarkers. AIDS 2017; 31 Suppl 2:S105–S119.

5. Rodés B, Cadiñanos J, Esteban-Cantos A, Rodríguez-Centeno J, Arribas JR. Ageing with HIV: Challenges and biomarkers. eBioMedicine 2022; 77. Available at: https://www.thelancet.com/journals/ebiom/article/PIIS2352-3964(22)00080-9/fulltext. Accessed 30 January 2025.

6. McHugh D, Durán I, Gil J. Senescence as a therapeutic target in cancer and age-related diseases. Nat Rev Drug Discov 2024; 24:57–71.

7. Cillo AR, Vagratian D, Bedison MA, et al. Improved single-copy assays for quantification of persistent HIV-1 viremia in patients on suppressive antiretroviral therapy. J Clin Microbiol 2014; 52:3944–3951.

8. Bruner KM, Wang Z, Simonetti FR, et al. A quantitative approach for measuring the reservoir of latent HIV-1 proviruses. Nature 2019; 566:120–125.

9. Rohart F, Gautier B, Singh A, Lê Cao K-A. mixOmics: An R package for ‘omics feature selection and multiple data integration. PLoS Comput Biol 2017; 13:e1005752.

10. Rohani R, Malakismail J, Njoku E. Pharmacological and Behavioral Interventions to Mitigate Premature Aging in Patients with HIV. Curr HIV/AIDS Rep 2023; 20:394–404.

11. Xing Y, Shepherd N, Lan J, et al. MMPs/TIMPs imbalances in the peripheral blood and cerebrospinal fluid are associated with the pathogenesis of HIV-1-associated neurocognitive disorders. Brain Behav Immun 2017; 65:161–172.

12. Copeland NK, Eller MA, Kim D, et al. Brief Report: Increased Inflammation and Liver Disease in HIV/HBV-Coinfected Individuals. J Acquir Immune Defic Syndr 2021; 88:310–313.

13. Chung NPY, Ou X, Khan KMF, Salit J, Kaner RJ, Crystal RG. HIV Reprograms Human Airway Basal Stem/Progenitor Cells to Acquire a Tissue-Destructive Phenotype. Cell Rep 2017; 19:1091–1100.

14. Ohene-Nyako M, Persons AL, Forsyth C, Keshavarzian A, Napier TC. Matrix Metalloproteinase-9 Signaling Regulates Colon Barrier Integrity in Models of HIV Infection. J Neuroimmune Pharmacol 2024; 19:57.

15. Brenchley JM, Price DA, Schacker TW, et al. Microbial translocation is a cause of systemic immune activation in chronic HIV infection. Nat Med 2006; 12:1365–1371.

16. Bassez A, Vos H, Van Dyck L, et al. A single-cell map of intratumoral changes during anti-PD1 treatment of patients with breast cancer. Nat Med 2021; 27:820–832.

17. Dhawan S, Wahl LM, Heredia A, et al. Interferon-γ inhibits HIV-induced invasiveness of monocytes. Journal of Leukocyte Biology 1995; 58:713–716.

18. Leng SX, Margolick JB. Aging, sex, inflammation, frailty, and CMV and HIV infections. Cellular Immunology 2020; 348:104024.

19. J Heath JD Grant M. The Immune Response Against Human Cytomegalovirus Links Cellular to Systemic Senescence. Cells 2020; 9:766.

20. Schnittman SR, Hunt PW. CMV and Persistent Immune Activation in HIV. Curr Opin HIV AIDS 2021; 16:168–176.

