## Supplementary material for "Senescence-related cytokine levels are associated with HIV-1 serostatus and persistence": Supplemental Materials.docx

**Supplementary Tables**

Supplementary Table 1

| Characteristics | PWoH (n = 10) | PWH (n=27) |
| --- | --- | --- |
| Age, years (IQR) | 59 (56, 59) | 59 (56, 65) |
| CD4 cell count, cells/mm^3^ (IQR) |  | 634 (548, 760) |
| CD4/CD8 ratio |  | 0.79 (0.57, 1.40) |
| Nadir CD4 cell count, cells/mm^3^ (IQR) |  | 272 (148, 456) |
| Viral suppression, n (%) |  | 27 (100) |
| ART, n (%) |  |  |
| ABC+3TC+DTG |  | 6 (22%) |
| FTC+TDF+ATV/r |  | 1 (3.7%) |
| AZT+3TC+IDV/r |  | 1 (3.7%) |
| FTC+TAF+BIC |  | 2 (7.4%) |
| DRV/c+DTG |  | 1 (3.7%) |
| FTC+TAF+EVG/c |  | 3 (11%) |
| DRV/r+ETV+RAL |  | 1 (3.7%) |
| FTC+TAF+DTG |  | 5 (19%) |
| DTG+RPV |  | 1 (3.7%) |
| FTC+TAF+RAL |  | 1 (3.7%) |
| FTC+TAF+RPV |  | 4 (15%) |
| LPV/r+RAL |  | 1 (3.7%) |
| IQR, interquartile range; ABC, abacavir; 3TC, lamivudine; DTG, dolutegravir; FTC, emtricitabine; TDF, tenofovir disoproxil fumarate; TAF, tenofovir alafenamide; ATV, atazanavir; /r, boosted by ritonavir; AZT, zidovudine; IDV, indinavir; BIC, bictegravir; DRV, darunavir; /c, boosted by cobicistat; EVG, elvitegravir; ETV, etravirine; RAL, raltegravir; RPV, rilpivirine; LPV, lopinavir. | | |

**Supplementary Figures**

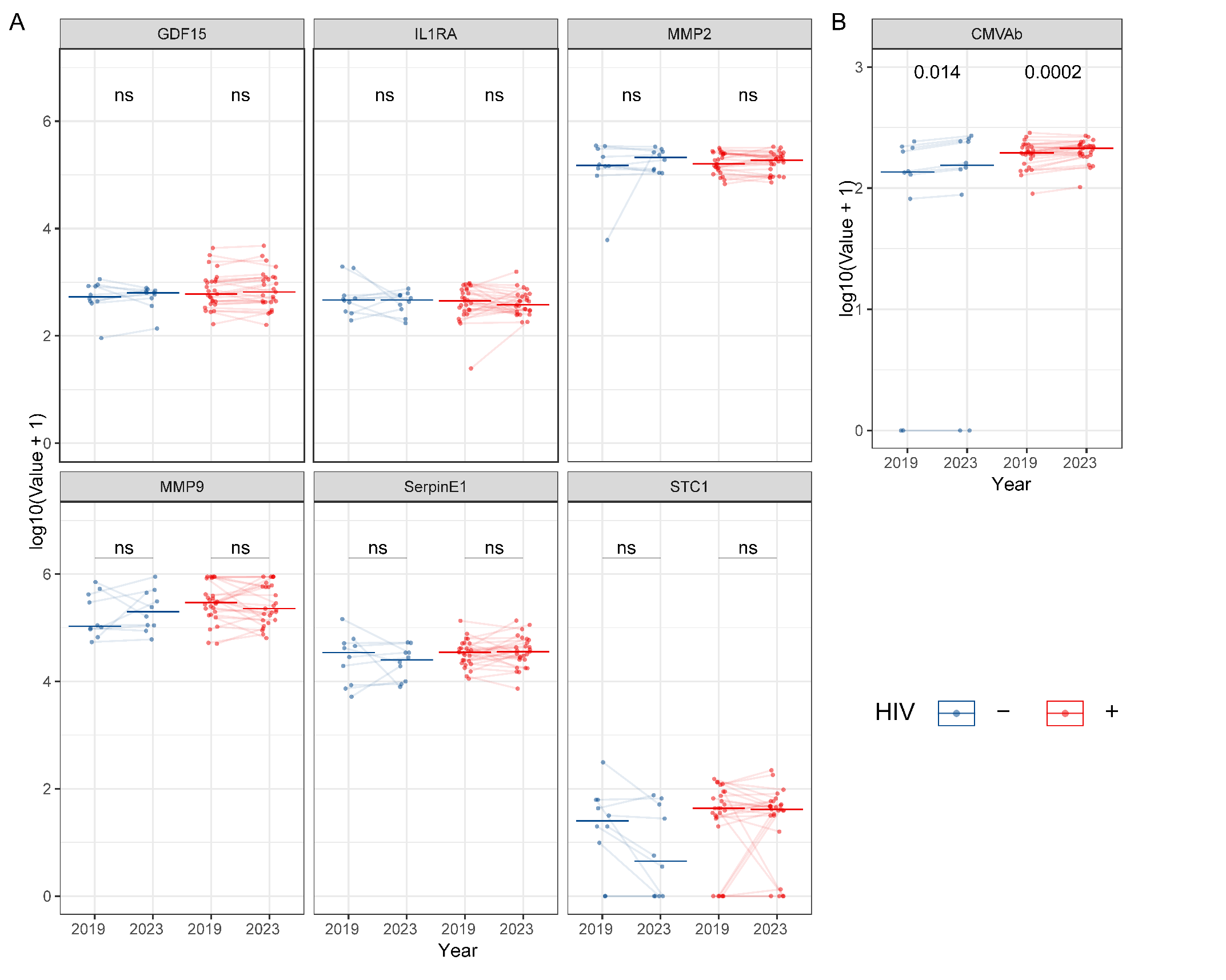

Supplementary Figure S1. SASP markers and CMVAb levels at years 2019 and 2023. (A) SASP markers. Units for SASP markers are all in pg/ml. (B) CMVAb levels (in relative unit [RU]/ml). ns, not significant.

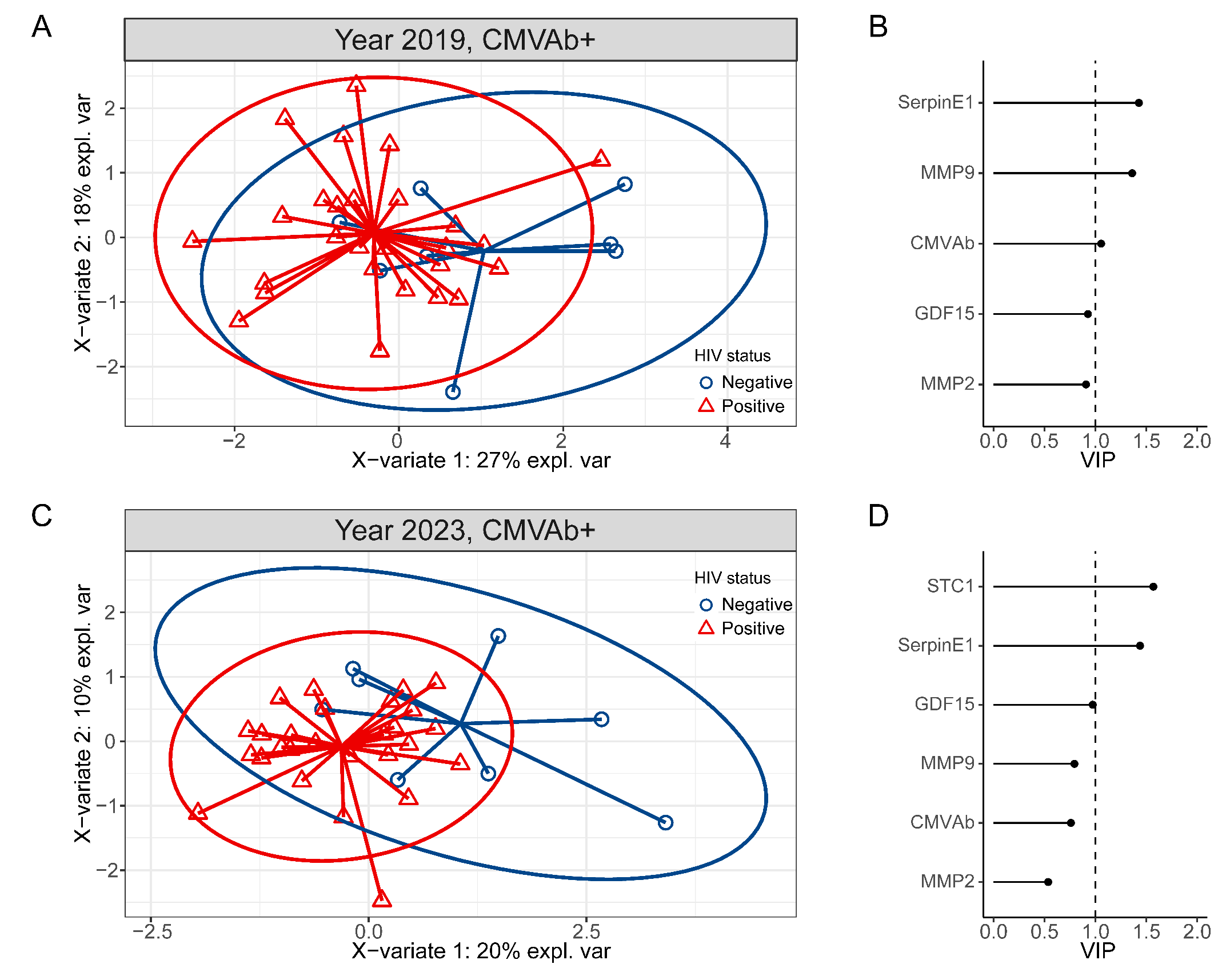

Supplementary Figure S2. Partial least squares discriminant analysis (PLS-DA) to evaluate the association between SASP markers, CMVAb and HIV serostatus in CMV seropositive participants. (A) SASP markers and CMVAb from 2019; (B) Variable importance in projection (VIP) of selected SASP markers from 2019 that can distinguish HIV serostatus. (C) SASP markers and CMVAb from 2019; (D) Variable importance in projection (VIP) of selected SASP markers from 2019 that can distinguish HIV serostatus.
